# Digital re-classification of equivocal dysplastic urothelial lesions using morphologic and immunohistologic analysis

**DOI:** 10.1101/2020.10.04.20206524

**Authors:** Camelia D Vrabie, Marius Gangal

## Abstract

A precise diagnostic of precursor dysplastic urothelial lesions is critical for patients but it can be a challenge for pathologists. Multiple immunohistologic markers (panel) improve ambiguous diagnostics but results are subjective, with a high degree of observational variability. Our research objective was to evaluate how a classification algorithm may help morphology diagnostic. Data coming from 45 unequivocal cases of flat urothelial lesions (“training set”: 20 carcinomas in situ, 8 dysplastic and 17 reactive lesions) were used as ground truth in training a random tree classification algorithm. 50 “atypia of unknown significance” diagnostics (diagnostic set) were digitally re-classified based on morphological and immunohistochemical features as possible carcinoma in situ (20), dysplastic (17) and reactive atypia cases (13). The main sorting criterium was morphologic (nuclear area). A four-markers panel was used for a precise classification (74% correctly classified, 93% accuracy, 76% precision, averaged ROC=0.828). 3 cases were “false negative”. The performance of the immunohistologic panel was evaluated based on a stain index, calculated for CD20, p53, Ki67 and observed for CD44. Within training set, the immunohistologic performance was high. In the diagnostic set both the percentage of high stain index for each marker and the percentage of cases with 2-3 strong markers were low, explaining the initial high number of equivocal cases. In conclusion, digital analysis of morphologic and immunohistologic features may bring clarification in classification of equivocal urothelial lesions. Computational pathology supports diagnostic process as it can measure features and handle data in a precise, reproducible and objective way. In our proof of concept study, a low number of cases and the (deliberate) absence of clinical data were main limitations. Validation of the method on a high number of cases, use of genomics and clinical data are essential for improving the reliability of machine learning classification

## Introduction

A precise diagnostic of flat urinary bladder lesions, critical for the treatment and prognosis of patients, can be difficult even for experienced uropathologists [1]. The most recent and widely used “flat urothelial lesions” classification (World Health Organization 2004 classification, modified in 2016) positioned Carcinoma in Situ (CIS), Dysplastic Lesions (DL), Reactive Hyperplasia (RH) and flat hyperplasia as part of a diagnostic continuum [2]. It is recommended to separate these diagnostic entities based on morphologic features [3] but a clear distinction is not always easy as bioptic tissue is scarce or fragmented, the normal urothelium histology is complex and important morphological features might be shared between normal, reactive and malign tissue [4]. CIS is described as a high-grade flat intraepithelial lesion composed of malignant cells with large hyper-chromatic nuclei (5-6x larger than a lymphocyte), serious pleomorphism and frequent mitoses, having a high propensity of developing muscle invasive cancers [5]. DL is tissue with recognizable pre-neoplastic cytological and architectural changes that are under the threshold of diagnostic of CIS [6]. RH shows smaller nuclei (2-3x the diameter of a lymphocyte nucleus), organized tissue structure, less nucleolar persistence [7] and is often associated with benign urologic pathology. A new diagnostic category, “Atypia of Unknown Significance” (AUS), sometimes named “urothelial proliferation of uncertain malignant potential” was recently added in an attempt to nest cases where a clear differentiation is difficult. AUS is still a subject of controversy and some authors recommend to avoid it as there is no robust evidence that patient outcomes are different from reactive atypia [8,9]. Because of overlapping morphology, a wide degree of subjectivity and inter-observational variability was reported mainly in equivocal cases [10,11].

Immunohistochemistry (IHC), a histology technique that allows a precise detection of specific proteins based on the antibody-antigen reaction, may improve precision when morphologic characteristics are not enough for a clear diagnostic, when the history of the disease is unknown or there is an unusual morphologic presentation [12]. After 20 years of experience, caution is now suggested in IHC interpretation of urothelial lesions [13] as normal tissue may share some degree of staining patterns and there is no single specific/sensitive marker that can make a difference between diagnostics. A way to circumvent the IHC ambiguities is provided by the concomitant use (panels) of several IHC markers [3]. Concomitant use of 4 markers (IHC panel composed of CK20, CD44, p53, and KI-67) demonstrated a clear adjuvant value to morphology [14-16]. CD20 (cytoplasmatic) is widely recognized as the most reliable CIS associated marker [17]. KI67 (nuclear) is seen as an aggressiveness and prognostic indicator [18], usually evaluated in connection with CD20 [19]. p53 (nuclear) is present in more than 50% of CIS cases and is often evaluated in conjunction with CD20 as well [20]. CD44 (membranous) is considered an exclusion marker [21,22]. Even with the help of IHC panels, a precise diagnostic is not always possible and many cases are still classified as AUS.

Computational pathology (CP) help improving the histologic exam accuracy [23]. Digital image analysis (DIA) can assess any type of microscopic image, including IHC [24]. DIA consists of image data acquisition and ground truth generation, image analysis (object detection, segmentation and recognition) and finally statistical evaluation of digital data [25]. When data is complex or equivocal, machine learning (ML) techniques can use mathematical models (algorithms) for statistical analysis. In the case of flat urothelial lesions diagnostic, CP can use an algorithm to analyze DIA data and generate associations in an objective, measurable and reproducible way. Our research objective was to evaluate how a decision tree algorithm can reclassify the AUS diagnostics after a supervised learning instruction process [26], using both morphology and IHC (4-panel) data.

## Material and methods

In this retrospective non-interventional analysis, precursor urothelial diagnostics found over a period of 3 years were re-evaluated by the principal investigator. All cases had a similar histology evaluation (Hematoxylin eosin and panel IHC) and were processed and stained using the same methodology (details concerning IHC characteristics and technique are provided in annex 1). Aside demographic info (sex and age), no other clinical information was available for this study.

Based on initial diagnostic classification, 2 datasets were created. An unequivocal “training” dataset (CIS, DL and RL) and equivocal “diagnostic” dataset (AUS). Images from both datasets were digitally analyzed in 2 steps: Data gathering (microscopic image capture, image segmentation, features measurement) was followed by data processing (data analysis, algorithm training and re-classification).

### Data gathering

Microscopic images were captured using standard microscopy (objective x20 and x40, camera 12MP) and stored on a desktop computer.

1. For morphology analysis, x20 images were manually segmented [27,28] under 3 labels (details in annex 1, table 2a). Probability maps were evaluated with the help of an open source software [29]. Binary images were used for digital measurement of nuclear area surface and roundness (length over width ratio) (image 1). The nuclear differentiation area threshold was based on previously published data [30,31]. Structural changes of layers and nuclear atypia were evaluated using a semiquantitative scale (yes, no), by direct observation on binary images.

**Image 1.**
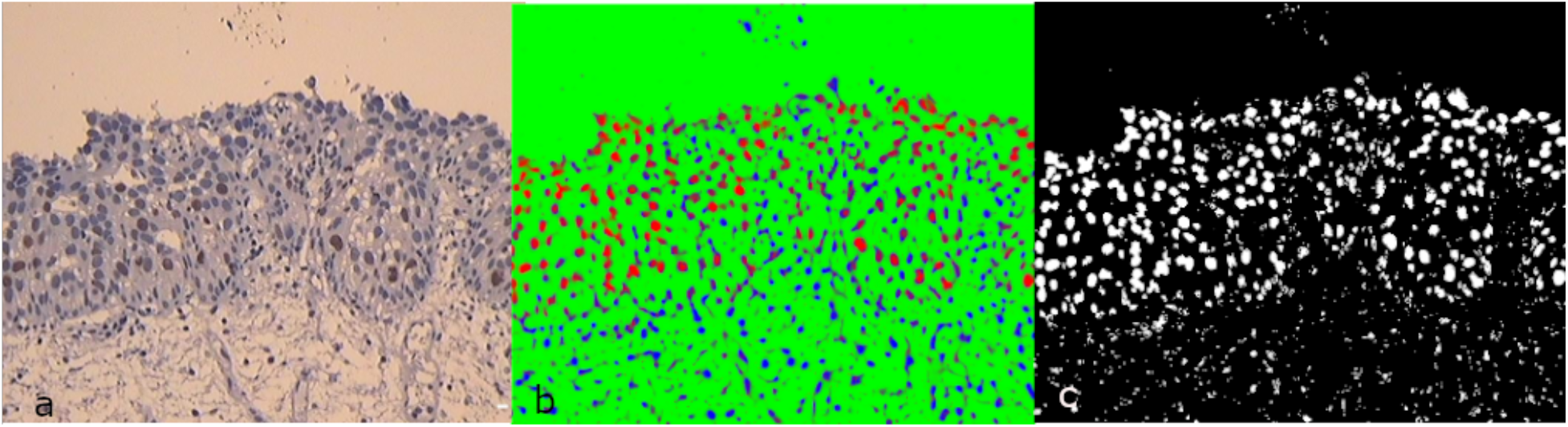
Morphology analysis. (a) IHC image, Ki67, x20. (b) Segmented image (3 labels). (c) Binary image. Atypia {yes}, Layer preservation {yes}, Bar: 40 microns
2. IHC markers evaluation was performed on x40 segmented images, in 2 steps. First, cytoplasmatic and nuclear markers images were segmented (3 or 4 labels). A Region of Interest (ROI) was delineated by human pathologist in an area of maximal stain intensity. An optical density index (ODI) was determined based on ROI histogram (image 2) [32]. Using a dedicated plugin (IHC profiler), the total stained surface was measured in terms of pixel intensity of DAB stain (image 3) [33]. Finally, an IHC stain index (SI) was calculated for each IHC marker (adding only high positive and positive pixels and multiplying the sum by ODI value).

**Image 2.**
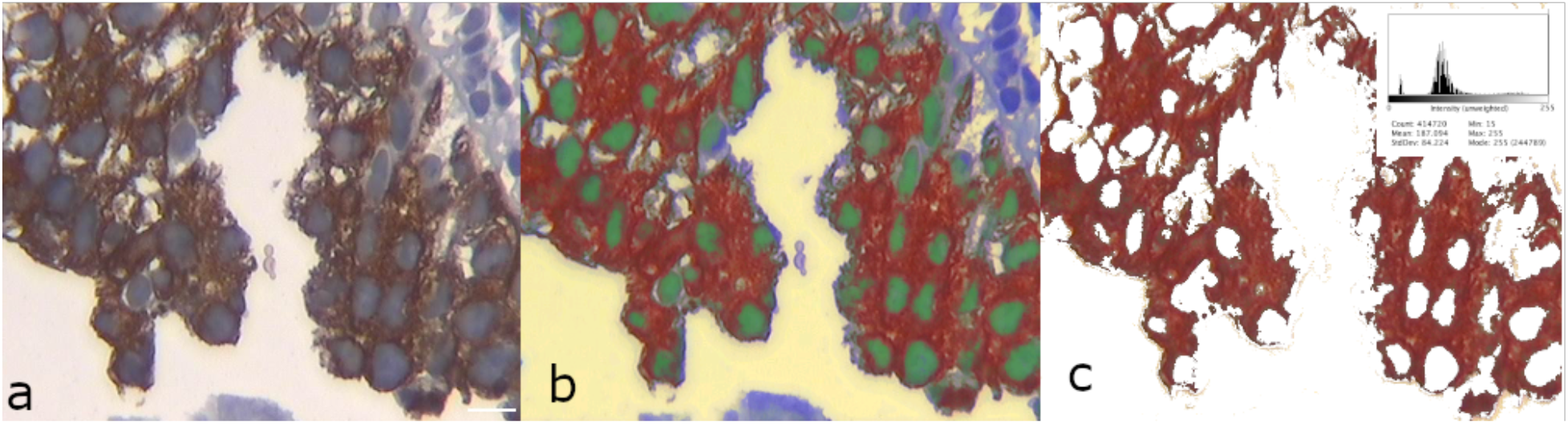
CD20 sampling process. (a) IHC stain x40. (b) Segmented image (4 labels). (c) Intensity of staining inside ROI: histogram. Bar: 40 microns

**Image 3.**
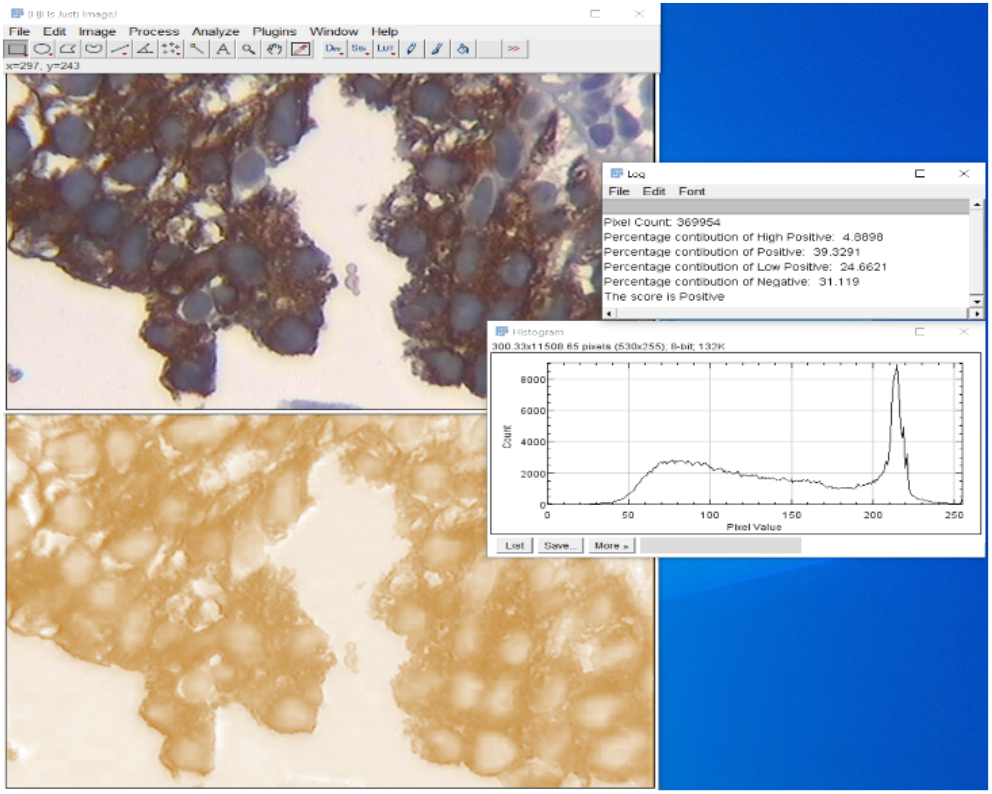
IHC index calculation. Cytoplasmatic marker surface measurement (%) with an open source software and a dedicated plugin (IHC profiler). Surface stained: 68.86%
3. CD44, as membranous marker, was measured semi-quantitatively (positive, absent or patchy).

### Data processing

A random decision tree classification algorithm was used for re-classification of all AUS cases. The “training” dataset (including all cases with a clear diagnostic of CIS, RH, DL and 5 cases of normal urothelium 50 instances) was considered ground truth and was used for algorithm training. The “diagnostic” dataset included all cases of equivocal atypia discovered in the clinic in the same period of time (50 instances).

As both datasets were small (100 instances, 16 attributes each) we opted for an open source machine learning toolkit (WEKA) software solution that is widely accepted in bioinformatics, has a good graphical interface and requires no programming [34]. A random tree classification algorithm was selected over the classic “c4.5” as graphical representation of the decision is as intuitive but cross validating classification results were better. Datasets attributes, quantification methodology and cross-validation results are detailed in annex 1. The structure of the random tree is presented in image 4.

**Image 4.**
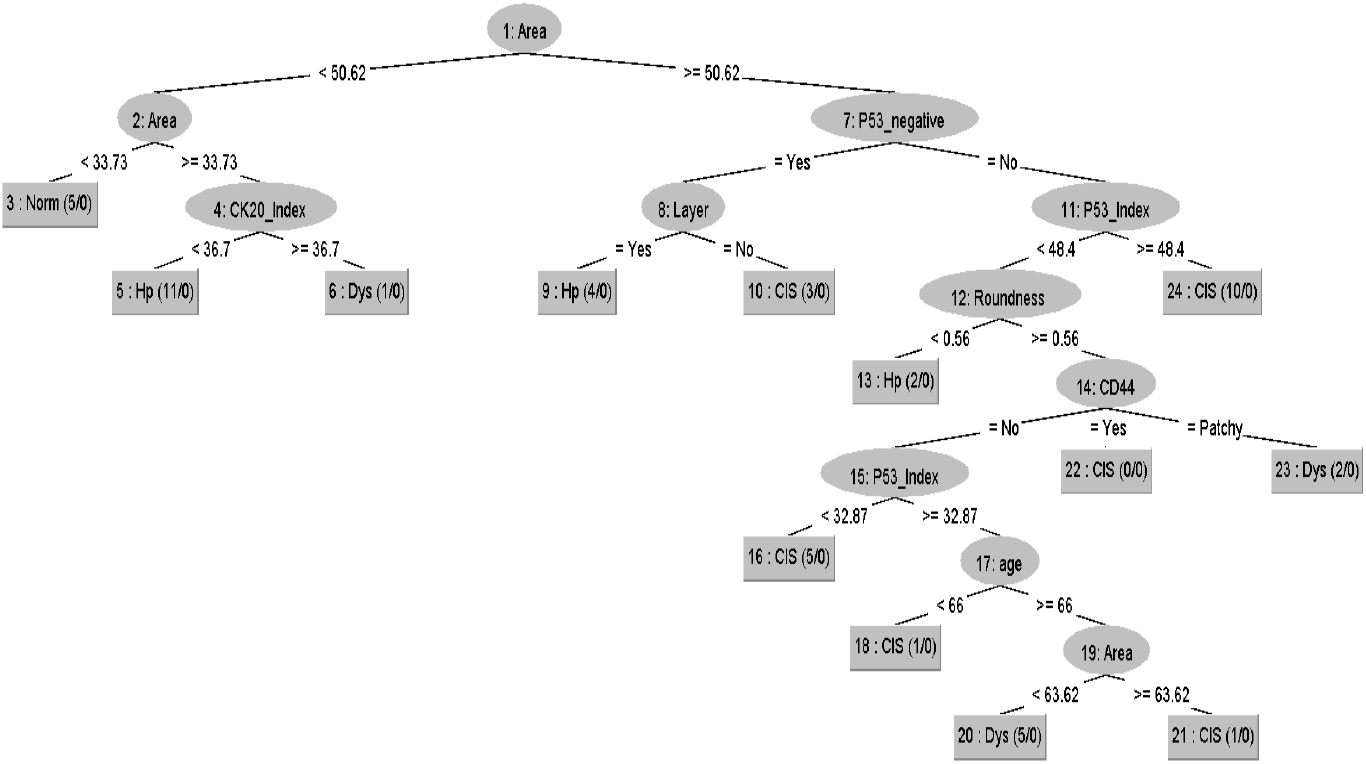
Decision tree algorithm, training dataset, classification criteria.

Based on intensity of stain for each marker, we recorded the percentage of positive cases in each group (SI threshold for CD20 and p53 > 50% and for Ki67 >15%). We also counted the concomitant presence of strong stain for each marker in neoplastic against non-neoplastic groups in both training and diagnostic sets.

### Statistics

All data was stored in a standard spreadsheet. Numerical data (age, nuclear diameter and nuclear area) was analyzed using MedCalc Statistical software (version 19.0.7, MedCalc Software bvba, Ostend, Belgium). Basic statistic results were expressed as mean ± standard deviation. Differences between means were tested for significance with a p-value set at p<0.05.

### Ethics

Patients provided written, informed consent before surgery specifically agreeing with the processing and analyze of the pathology urothelium samples. The retrospective, non interventional IHC study protocol was approved by the hospital IRB (1123/2020).

## Results

From 256 existing urinary bladder biopsies over 3 years (188 men, 68 women), 191 neoplastic cases were selected. 71 of cases received a diagnosis of T1 and 18 of T2 muscular infiltrative carcinomas. 20 cases were classified as CIS, 8 cases were DL and 17 HL based on unequivocal morphologic and IHC features. 50 cases with an initial AUS diagnostic, were re-classified based on ground truth morphologic and IHC characteristics in 20 CIS, 17 DL and 13 RH. Results are summarized in table 1 (training dataset) and table 2 (diagnostic dataset).

**Table 1.** Training dataset measurement results (*p<0.05, **p<0.0001 when compared to CIS nuclear area) (CD44: y=yes, pt=patchy, n=no)

|  | Training set |  |  |  |
| --- | --- | --- | --- | --- |
|  | CIS | DL | HL | Normal |
| **Nr.** | 20 | 8 | 17 | 5 |
| **Sex** | 1f/19m | 0f/8m | 3f/14m | 2f/3m |
| **Age** | 73.75+6.7 | 73.38+6.5 | 67.82+8.3 | 63.4+10.6 |
| **Nuclear Area ($\mu^2$)** | 59.42+4.9 | 54.54+0.9* | 49.09+5.4** | 32.47+1.4 |
| **Layer preservation** | 20% | 37.50% | 75% | 100% |
| **Nuclear Roundness** | 0.59+0.02 | 0.57+0.02 | 0.54+0.02 | 0.49+0.01 |
| **Cell Atypia** | 95% | 35% | 17.60% | 0% |
| **CK20 Superficial** | 5% | 12.50% | 58.80% | 80% |
| **CK20 SI** | 49.69+11.7 | 44.3+13.9 | 9.56+9.4 | 1.24+0.43 |
| **p53 Negative** | 10% | 12.50% | 58.80% | 100% |
| **p53 SI** | 37.02+17.5 | 38.53+13.4 | 17.00+12.6 | 3.45+1.3 |
| **Ki67 Negative** | 10% | 12.50% | 53% | 80% |
| **Ki67 SI** | 14.36+5.1 | 10.65+3.06 | 7.58+1.8 | 4.52+2.9 |
| **CD44** | 0%y/5%pt/95%n | 0%/25%pt/75%n | 83%y/17%pt/0%n | 75%y/25%pt/0%n |

**Table 2.** Diagnostic dataset. Measurement results after reclassification (*p<0.05, **p<0.0001 when compared to CIS nuclear area) (CD44: y=yes, pt=patchy, n=no)

|  | Diagnostic set |  |  |
| --- | --- | --- | --- |
|  | CIS | DL | HL |
| **No** | 20 | 17 | 13 |
| **Sex** | 2f/18m | 2f/15m | 0f/13m |
| **Age** | 71.05+5.6 | 72.71+7.3 | 69.61+7.9 |
| **Nuclear Area ($\mu^2$)** | 57.95+5.02 | 50.72+5.36** | 45.25+4.3** |
| **Layer preservation** | 40% | 17% | 46% |
| **Nuclear Roundness** | 0.59+0.03 | 0.58+0.03 | 0.57+0.04 |
| **Cell Atypia** | 35% | 47% | 30.70% |
| **CK20 Superficial** | 0% | 0% | 38.50% |
| **CK SI** | 38.64+13.9 | 43.16+9.37 | 19.82+14.4 |
| **p53 Negative** | 35% | 11.70% | 38.50% |
| **p53 SI** | 26.1+19.8 | 29.04+13.1 | 21.27+16.61 |
| **Ki67 Negative** | 25% | 35.25% | 30.70% |
| **Ki67 SI** | 8.53+4.66 | 7.35+5.04 | 7.051+3.9 |
| **CD44** | 0%y/0%pt/100%n | 0%y/11%pt/89%n | 0%y/46%pt/54%n |

## Discussion

Urothelial carcinoma is the 10th most common and the second most frequent genitourinary malignancy, worldwide [35]. As urothelial carcinoma has a high recurrence rate, an early, precise differentiation of precursor neoplasms from benign reactive atypia is critical [36]. Because many flat lesions morphologic and IHC features may overlap or are shared by normal urothelium, the diagnostic is sometimes equivocal with a possible increase of medico-legal risk [37].

Our research objective was to evaluate how a machine learning algorithm can reclassify ambiguous precursor flat lesions diagnostics that were previously included under the AUS diagnostic category. A random decision tree classification algorithm was trained using data from all unequivocal flat lesions diagnosed in our unit over 3 years (20 CIS, 17 HL, 8 DL). 5 normal urothelium IHC stained tissue were also included in the training dataset for classification purposes only. The training process used both morphologic and a “classic” 4-markers IHC panel data.

Both datasets (training and a diagnostic) were digitally evaluated using open label software and affordable computational technology.

Based on classification information provided in the training dataset, the random tree algorithm reclassified the diagnostic dataset (50 AUS cases). 20 CIS, 17 DL and 13 HL possible diagnostics were suggested.

In the classification process, as expected, the nuclear area played a central role in differentiating precursor malign from benign diagnostics. Further, layer preservation, circularity and IHC calculated index were used at different levels of classification. Reported differences between the nuclear area measured in diagnostic set were by 9.7% smaller than in the training set (p=0.35), making human optical measurement and differentiation difficult. Similar difficulties were noticed for all other morphologic classification criteria.

The specific performance of the IHC panel was analyze based on percentage of strong SI for each marker in each set. Results are presented in table 4. In the training set, CIS/DL group had a 3-markers strong positivity in 23.75% of cases and of 2-markers in 56.25% of cases. In the diagnostic dataset only 7.5% of cases had 3-markers and 27% a 2-markers strong presence. For HL patients, results were negative for all inclusion markers except Ki67 in 5 cases. All cases in the diagnostic dataset were CD44 negative (13 cases were considered “patchy”).

**Table 3.** Confusion matrix, Random tree classification, training data set, (*) false negative cases meaning malign cases classified as benign or benign as malign. Mis-classification within same classes were not penalized (CIS as DL or HL as normal)

| **Confusion matrix** |  |  |  |  |  |
| --- | --- | --- | --- | --- | --- |
| **a** | **b** | **c** | **d** | **e** | **Classified as:** |
| **14** | **1\*** | 0 | 5 | 0 | a = CIS |
| **0** | 14 | 0 | **1\*** | 2 | b = HL |
| **0** | 0 | 0 | 0 | 0 | c = AUS |
| **2** | **1\*** | 0 | 5 | 0 | d = DL |
| **0** | 1 | 0 | 0 | 4 | e = Norm |

**Table 4.**
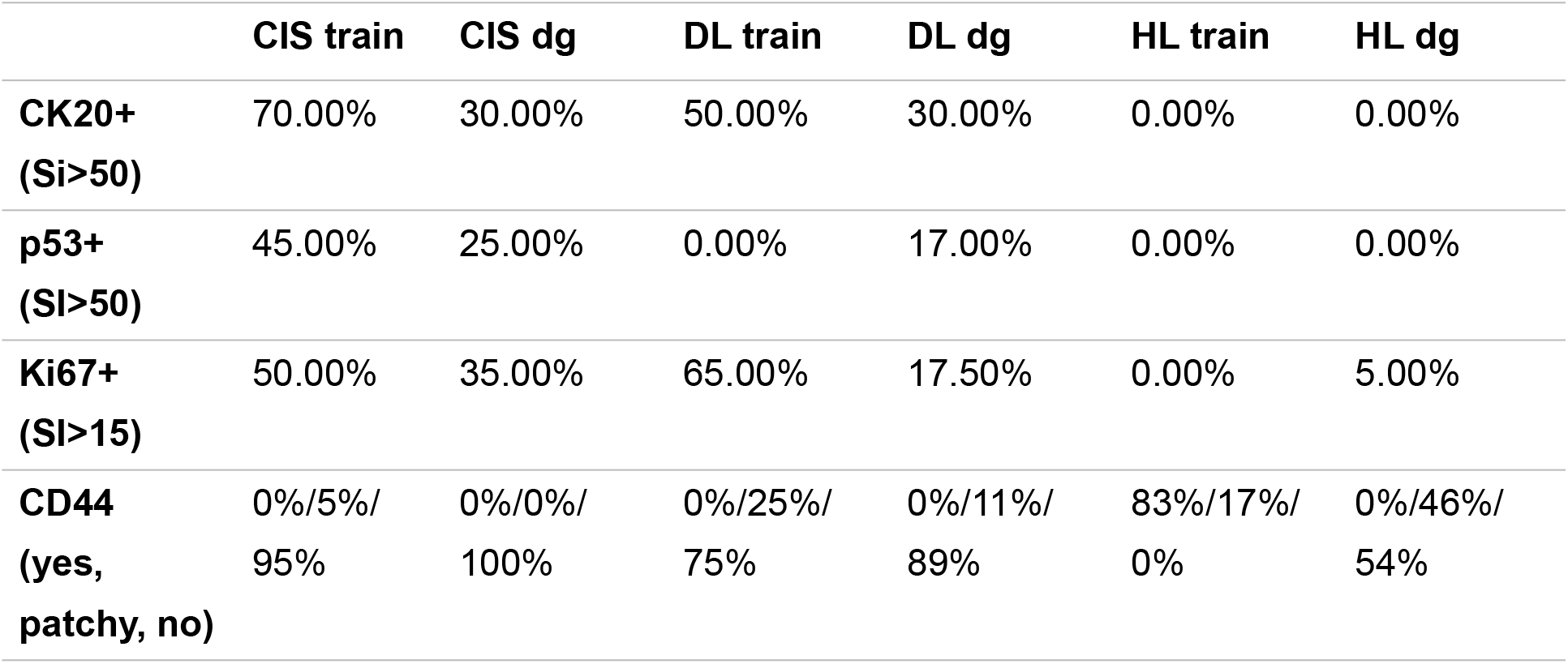
IHC panel performance based on staining intensity for different diagnostic groups in training and diagnostic datasets (after ML re-classification)

Overall, the random tree algorithm classification had a good performance (74% correctly classified, 93% accuracy, 76% precision) when measured against the ground truth. The receiver operating characteristics (ROC) for CIS was 0.817, for DL was 0.741 and for HL 0.866. Important, only 3 cases were “false negative” (one CIS and one DL cases were classified as HL and one HL as DL) (confusion matrix table 3).

In conclusion, urothelial flat lesion classification can be a diagnostic challenge on bioptic material. Usually, a distinction between malign and benign cases is made using morphologic features and IHC is used mainly for confirmatory reasons. Both diagnostic methods are subjective and may have inter and intra observational variability and may generate ambiguous diagnostics. Our objective was to evaluate how CP can be used to circumvent flat lesions diagnostics uncertainty. Digital data from 45 unequivocal urothelial flat lesions data were used for training a ML algorithm (random tree). Main criteria for algorithm classification were morphological and a calculated IHC stain index coming from a classical panel of 4 IHC markers. The performance of the classification algorithm was good: 93% accuracy and 76% precision, averaged ROC 0.828. Only 3 cases were classified as “false negative”. Based on this “ground truth”, 50 equivocal diagnostics (AUS) were reclassified as 40% CIS, 34% DL and 26% HL. The IHC panel performance in classification was finally judged based on each of the 4 markers SI: in the training set, the panel performed well but in the diagnostic set, the performance was overall low.

Main limitations of this study are the low number of analysed cases (explaining the 76% precision rate), the deliberate absence of clinical data, absence of genomic information and the use of a non-customizable classification algorithm. The number of flat urothelial lesions is low and reflects the rarity of this diagnostic in everyday uro-pathology practice. The absence of clinical data is an important clinical limitation as many of flat lesions are recurrent. Long term clinical follow-up of cases, use of a molecular phenotyping information and a customized classification algorithm may further improve classification precision.

In flat atypia equivocal cases, CP can be a support for diagnostics, as it may handle multiple data attributes in a fast, effective and precise way.

## Data Availability

All information is provided in the article and in the Annex

## Annex

**Table 1a.**
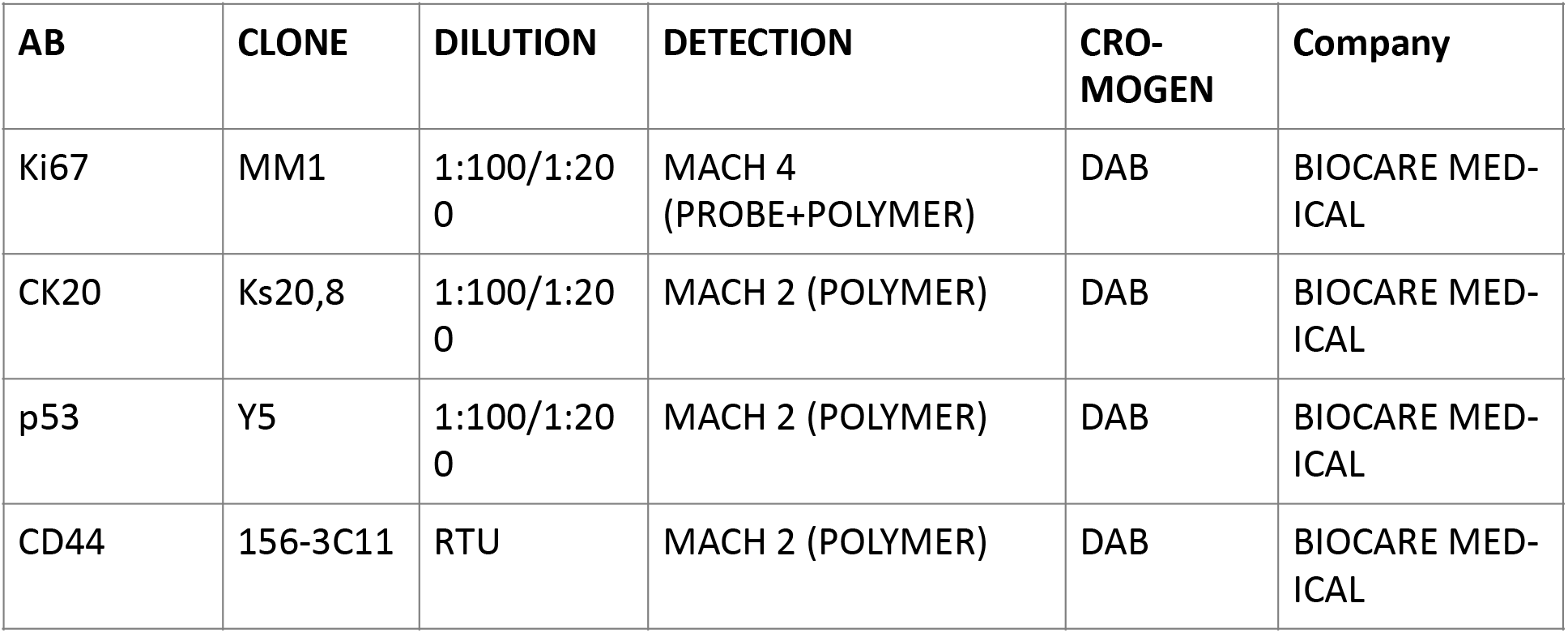
IHC Markers Characteristics. Histologic staining was performed on 4-µm-thick sections from formalin-fixed, paraffin-embedded tissue blocks.

| AB | CLONE | DILUTION | DETECTION | CRO-MOGEN | Company |
| --- | --- | --- | --- | --- | --- |
| Ki67 | MM1 | 1:100/1:200 | MACH 4 (PROBE+POLYMER) | DAB | BIOCARE MEDICAL |
| CK20 | Ks20,8 | 1:100/1:200 | MACH 2 (POLYMER) | DAB | BIOCARE MEDICAL |
| p53 | Y5 | 1:100/1:200 | MACH 2 (POLYMER) | DAB | BIOCARE MEDICAL |
| CD44 | 156-3C11 | RTU | MACH 2 (POLYMER) | DAB | BIOCARE MEDICAL |

**Table 2a.** IHC Segmentation labels

|  |  |
| --- | --- |
| Segmentation x20 | IHC atypia cells, normal cells, background |
| Segmentation x40 | Cytoplasm, atypia nucleus, normal nucleus, background |

**Table 3a.**
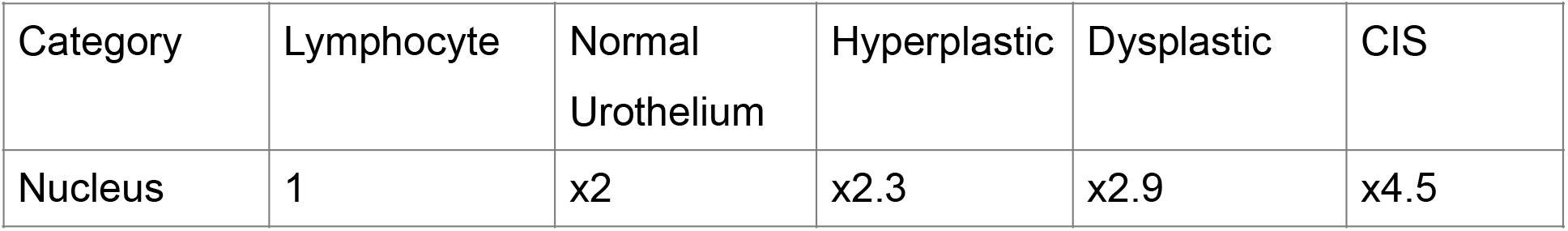
Morphologic threshold for DIA measurements in different flat urothelial lesions

| Category | Lymphocyte | Normal Urothelium | Hyperplastic | Dysplastic | CIS |
| --- | --- | --- | --- | --- | --- |
| Nucleus | 1 | x2 | x2.3 | x2.9 | x4.5 |

**Table 4a.** Attributes and quantification methods used in algorithm training and classification datasets (CIS: carcinoma in situ, Hp: Reactive Hyperplasia, AUS: atypia of unknown significance, Dys: dysplasia, Norm: Normal urothelium)

| Attribute | Quantification |
| --- | --- |
| Sex | {f, m} |
| Age | numeric |
| Area | numeric |
| Structure | {Yes, No,} |
| Circularity | Numeric |
| Atypia | {0, 1, 2} |
| CK20_superficial | {Yes, No} |
| CK20_index | numeric |
| p53 _negative | {Yes, No} |
| p53 _index | numeric |
| Ki67 _negative | {Yes, No} |
| Ki67_index | numeric |
| CD44 | {No, Yes, Patchy} |
| Dg | {CIS, Hp, AUS, DL, Norm} |

Details of classification accuracy – training set

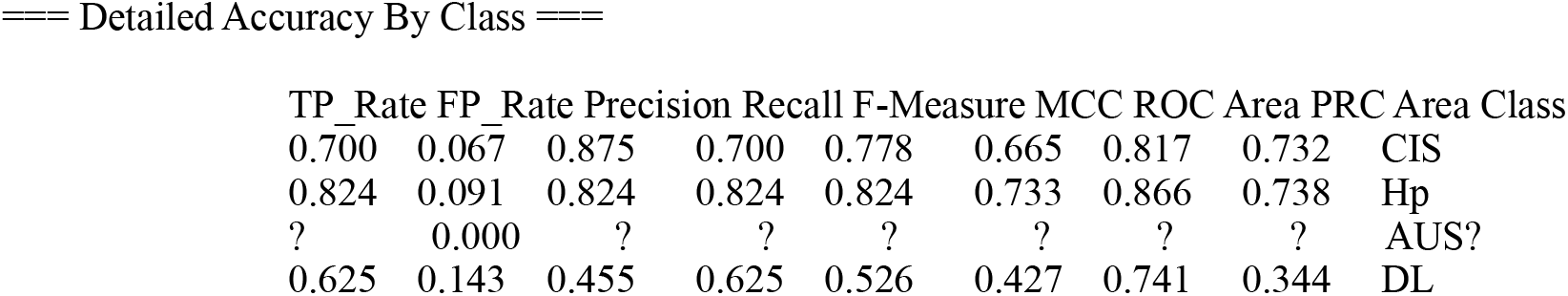
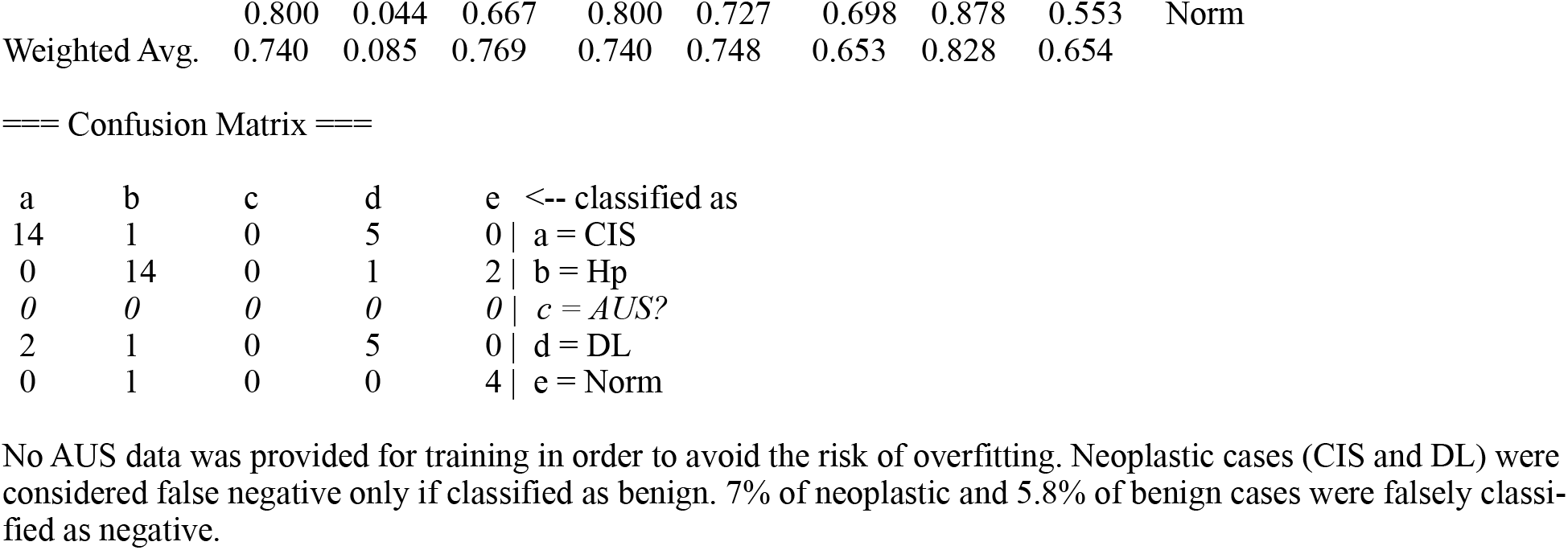

| TP_Rate | FP_Rate | Precision | Recall | F-Measure | MCC | ROC Area | PRC Area | Class |
| --- | --- | --- | --- | --- | --- | --- | --- | --- |
| 0.700 | 0.067 | 0.875 | 0.700 | 0.778 | 0.665 | 0.817 | 0.732 | CIS |
| 0.824 | 0.091 | 0.824 | 0.824 | 0.824 | 0.733 | 0.866 | 0.738 | Hp |
| ? | 0.000 | ? | ? | ? | ? | ? | ? | AUS? |
| 0.625 | 0.143 | 0.455 | 0.625 | 0.526 | 0.427 | 0.741 | 0.344 | DL |

|  |  |  |  |  |  |  |  |  |  |
| --- | --- | --- | --- | --- | --- | --- | --- | --- | --- |
|  | 0.800 | 0.044 | 0.667 | 0.800 | 0.727 | 0.698 | 0.878 | 0.553 | Norm |
| Weighted Avg. | 0.740 | 0.085 | 0.769 | 0.740 | 0.748 | 0.653 | 0.828 | 0.654 |  |

=== Confusion Matrix ===
|  |  |  |  |  |  |
| --- | --- | --- | --- | --- | --- |
| a | b | c | d | e | <-- classified as |
| 14 | 1 | 0 | 5 | 0 | a = CIS |
| 0 | 14 | 0 | 1 | 2 | b = Hp |
| 0 | 0 | 0 | 0 | 0 | c = AUS? |
| 2 | 1 | 0 | 5 | 0 | d = DL |
| 0 | 1 | 0 | 0 | 4 | e = Norm |

No AUS data was provided for training in order to avoid the risk of overfitting. Neoplastic cases (CIS and DL) were considered false negative only if classified as benign. 7% of neoplastic and 5.8% of benign cases were falsely classified as negative.

